# COVID-19 SCREENING AND MONITORING OF ASYMPTOMATIC HEALTH WORKERS WITH A RAPID SEROLOGICAL TEST

**DOI:** 10.1101/2020.05.05.20086017

**Authors:** Angelo Virgilio Paradiso, Simona De Summa, Nicola Silvestris, Stefania Tommasi, Antonio Tufaro, Giuseppe De Palma, Angela Maria Vittoria Larocca, Vincenzo D’Addabbo, Donata Raffaele, Vito Cafagna, Vito Garrisi

**Affiliations:** Scientific Directorate, IRCCS-Istituto Tumori “Giovanni Paolo II”, Via O.Flacco, 65-70124 Bari, Italy; Molecular Diagnostics and Pharmacogenetics Unit, IRCCS-Istituto Tumori “Giovanni Paolo II”, Via O.Flacco, 65-70124 Bari, Italy; Department of Biomedical Sciences and Human Oncology, Unit of Internal Medicine “Guido Baccelli”, University of Bari Medical School, 70124 Bari, Italy; MedicalOncology Unit, IRCCS Istituto Tumori “Giovanni Paolo II” of Bari, Viale Orazio Flacco, 65, 70124 Bari, Italy; InstitutionalBioBank, ExperimentalOncology and Biobank Management Unit, IRCCSIstituto Tumori “Giovanni Paolo II”, Via O.Flacco, 65-70124 Bari, Italy; Hygiene Unit, Policlinico Hospital, Piazza G. Cesare 11, 70124 Bari, Italy; UOSSD Servizio Professioni Sanitarie, IRCCS-Istituto Tumori “Giovanni Paolo II”, Via O.Flacco, 6570124 Bari, Italy; Clinical PathologyLaboratory, IRCCS-Istituto Tumori “Giovanni Paolo II”, Via O.Flacco, 65-70124 Bari, Italy

## Abstract

**Background:** Health workers are at high risk for SARS-CoV-2 infection and, if asymptomatic, for transmitting the virus on to fragile cancer patients.

**Materials and method:** We monitored health care workers (HCW) of our Cancer Institute with the rapid serological test Viva-Diag^TM^ analyzing COVID-19 associated-IgG/IgM. Test were performed at time 0 and after 14 days; Rt-PCR and CLIA assays were also perfoRmed in positive Viva-Diag^TM^ cases. 606 and 393 HCW had blood sample taken at time 0 and 14, respectively.

**Results:** Overall, 9 HCW (1.5%) resulted not-negative at Viva-Diag^TM^ and one of them was confirmed positive for SARS-COV2 infection at RT-PCR oropharingeal swab. At time 0, all 9 cases showed some IgM expression and only one IgG; after 14 days IgM persisted in all cases while IgG became evident in 4 ones. A parallel CLIA test was performed in 23 quaratined subjetcs and in all Viva-Diag not negative cases. CLIA confirmed a positive level of IgM in 5/13 positive Viva-Diag cases; conversely, IgG was confirmed positive at CLIA in 4/5 cases positive at Viva-Diag. These results pose the question of different performances of the two tests.

**Conclusions:** Our study suggest that Viva-Diag assay can be of help in individualizing SARS_COV2 infected people fisrt of all in cohorts of subjetcs with high prevalence. Different performances of serological colorimetric and CLIA tools remain to be ascertained.

## Introduction

As of 25 Aprile 2020 there are 3,073,603 coronavirus disease 2019 (COVID-19) positive cases (211,768 deaths, 1,937,184 active cases with 1,880,893 in mild conditions and 56,291 in serious or critical conditions) (1). This pandemic storm has forced each of us into a life-threatening situation, with health systems, even in the most advanced countries, in crisis (2).

In this scenario, since the first cases have been reported in China, we have learned that this infection can present with very heterogeneous clinical pictures in terms of clinical severity, from completely asymptomatic to serious and potentially fatal forms (3). While the latter represent an often dramatic challenge, requiring in some cases hospitalization in intensive care units, on the other hand, asymptomatic forms represent one of the determining factors for the spread of the infection (4). This event assumes particular relevance for some categories of workers in particular health care workers (HCWs) considering their contacts with subjects who are frequently frail due to the pathologies from which they are affected and for which they turn to them. In particular, cancer patients represent a risk category with significant management challenges associated to both the risk of contagion and the possible major complications related to the infection (5–6). The incidence of contracting COVID-19 infection has been 16% for HWs who carried out their activity in departments dedicated to COVID-19 patients (7). This percentage decreased to 4.1% for HCWs who worked in wards with patients with respiratory symptoms without laboratory evidence of positivity for the virus (8). To date there are no published data indicating the incidence of asymptomatic positive COVID-19 HCWs working at a center dedicated exclusively to the treatment of cancer patients.

RT-PCR on naso-pharyngeal swabs is the standard test although it presents sensitivity problems and is not easily applicable to the general population. Serological tests, rapid and easy to handle, permit to study immunoglobulin expression of a virus contact. In particular, it is of particular interest the analysis of IgM and IgG which express recent or late contact with the virus (9). Nevertheless, this data acquired during previous Coronavirus epidemic diseases, is still to be defined for COVID-19 (10).

For these reasons, we planned to screen a cohort of HWs at Istituto Tumori “Giovanni Paolo II” of Bari (National cancer research Center) to verify the following issues: 1) the applicability of a rapid serological test to an asymptomatic HCW population; 2) the prevalence of SARS-CoV2 immunoreaction; 3) the kinetics if IgM/IgG in HCWs at 2 weeks interval; 4) the comparison of rapid serological test results with respect to RT-PCR and CLIA assay

## Material and methods

From March 26th 2020, all asymptomatic HCWs employed at Istituto Tumori G. Paolo II, IRCCS, of Bari were invited to take part in a prospective trial planning to monitor SARS-CoV2 serological profile (IgM/IgG) in venous blood sample by the rapid serological test, VivaDiag^TM^. Blood samples were planned to be taken upon entering the study and repeated after 2 weeks. Furthermore, the protocol planned: to perform standard RT-PCR assays carried out on oropharyngeal swabs of people with Viva-Diag^TM^ test providing not-negative IgM/IgG result; in a subset of cases, the comparison between Viva-Diag^TM^ results and IgM/IgG analysis perfomed by the Chemio-Luminescence Assay (CLIA) MAGLUMI800™ was done, also.The study was approved by the Ethical Committee of the IstitutoTumori G. Paolo II, IRCCS, Bari with Protocol number CE 872/2020.

A total of 606 health workers (94% of the total population entered the first round of the study while 393 (65%) completed the monitoring phase (with the second blood sample) at time of study closing (April, 17th). After signing the written informed consent, all participants filled out a questionnaire collecting information on presence of clinical symptoms and their possible risk of COVID-19 infection (contact with confirmed SARS-CoV2 positive individuals or visits to areas with active SARS-CoV-2 circulation). After collection, venous blood sample collection, was immediately sent to Clinical Pathology Laboratory (Certified ISO-9001/2015) and to the Institutional Biobank (Certified ISO-9001/2015) of the Istituto Tumori G Paolo II, IRCCS, Bari (I) to performr Viva-Diag^TM^ test.

The characteristics of the health workers who entered the study are reported in Table 1. Their median age was 47.5 years (range 20-73 yrs) and 39.4% were male. A total of 54.1% of the enrolled workers were involved in direct clinical activities, 9% in laboratory practice, 8% in administrative activities and 28.9% in maintenance/cleaning activities. 1.1% of them reported minor clinical symptoms not directly referable to COVID-19 disease while 11.7% reported having had direct contact with individuals with suspected COVID-19 disease in the last two weeks. Lastly, 6.7% of subjetcs returned to hospital activities after a quarantine period (none with previous SARS-CoV2 positive RT-PCR test). Operators who took part to the second part of the study did not statistically for clinical characteristics from the whole starting series

**Table 1.** Characteristics of the cohort of health workers screened for SARS-CoV-2

|  | **FIRST ROUND** | **SECOND ROUND** |
| --- | --- | --- |
| ***COHORT CHARACTERISTICS*** | **N=606 (%)** | **N=393 (%)** |
| ***Sex*** |  |  |
| Male | 239 (39.4%) | 142 (36.1%) |
| Female | 367 (60.5%) | 251 (63.9%) |
| ***Age*** | 47.49 years (range: 20-73) | 48.3 years (range: 20-66) |
| ***Role*** |  |  |
| Clinical activity | 328 (54.1%) | 212 (53.9%) |
| Laboratory | 54 (9%) | 36 (9.1%) |
| Administrative | 49 (8%) | 79 (20.1%) |
| Maintenance/cleaning | 175 (28.9%) | 66 (16.8%) |
| ***Subjects with SARS-CoV-2 contacts*** | 71 (11.7%) | 42 (10.7%) |
| ***Subjects with minor symptoms*** | 7 (1.1%) | 5 (1.2%) |
| ***Quarantined subjects*** | 41 (6.7%) | 23 (5.8%) |

### SARS-CoV-2 rapid IgG-IgM Test

SARS-CoV-2 rapid IgG-IgM combined antibody test kit, Viva-Diag^TM^, was designed and manufactured by Viva-Check BIOTECH (VivaChek Biotech, Hangzhou) is a lateral flow qualitative immunoassay for the rapid determination of the presence of both anti-SARS-CoV-2-IgM and anti-SARS-CoV-2-IgG in blood. A surface antigen from SARS-CoV-2 which can specifically bind to SARS-CoV-2 antibodies (including both IgM and IgG) is conjugated to colloidal gold nanoparticles and sprayed onto conjugate pads. The presence of SARS-CoV-2 IgG and IgM antibodies is indicated by a red/purple line that appears in the specific region for those antibodies on the device. Colorimetric reaction for IgM and IgG was separately evaluated and classified as Negative, with Weak or Strong colorimetric reaction. Each test was evaluated by two operators and a picture of the colorimetric result was taken. In case of disagreement between two operators, the picture was evaluated by a third party.

### Molecular detection of SARS-CoV-2

In presence of Viva-Diag^TM^test not-negative for IgM and/or IgG (n=9), oropharyngeal swabs were collected on the following day for standard SARS-CoV-2 RT-PCR testing. The RT-PCR tests were immediately performed at the Laboratory of Molecular Epidemiology and Public Health (Head:M.Chironna) of the University of Bari (I). Nucleic acid was extracted from swabs by MagNA Pure (Roche Diagnostics, Mannheim, Germany), according to the manufacturer’s instructions. The presence of the E gene, RdRP gene and N gene of the SARS-CoV-2 virus were identified by a commercial real-time PCR assay (Allplex 2019-nCoV Assay; Seegene, Seoul, Republic of Korea). Samples were considered positive if all the three genes were molecularly detected. The WHO Real-time-PCR protocol was used to confirm the presence of SARS-CoV2. (https://www.who.int/docs/default-source/coronaviruse/uscdcrt-pcr-panel-for-detection-instructions.pdf?sfvrsn=3aa07934_2).

### Chemiluminescence (CLIA) IgG/IgM detection

The IgG/IgM dosage was determined utilizing MAGLUMI800™ 2019-nCoV IgG (Cat. Ref. 130219015M) andIgM (130219016M) (CLIA) according to the manufacturer’s indications (Shenzhen New Industries Biomedical Engineering Co., Ltd; www.snibe.com). Briefly, the kit permits the indirect chemiluminescence qualitative-semiquantitative in vitro immunoassay of IgG and IgM antibodies against SARS-CoV-2in human blood.The pre-diluted biological sample, buffer and magnetic microbeads coated with recombinant SARS-CoV-2 antigen were mixed and incubated to create immunocomplexes. After magnetic field exposure IgG and IgM antibodies labeled with ABEI were added; the chemiluminescence starter activated the light reaction which was detectedby the photomultipier MAGLUMI800™ (www.snibe.com). Results were expressed as Relative Light Units (RLU) and considered positive if the signal/cutoff (S/C) ratio was ≥ 1.

### Statistical analyses

A descriptive analysis of the VivaDiag^TM^test results with respect to the health workers’ characteristics is shown in Table 1.VivaDiag^TM^ test results are compared to the other laboratory assay results in Table 2.

**Table 2.**
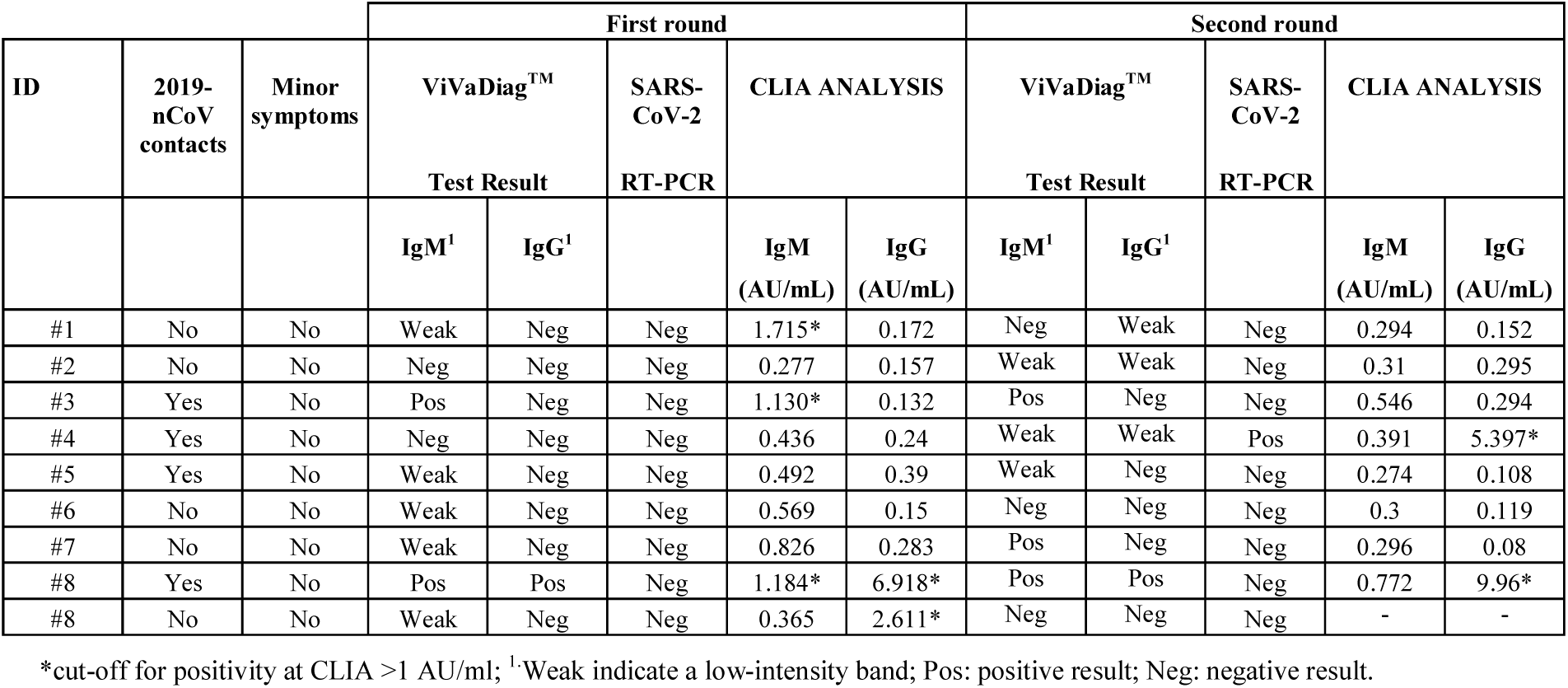
Results of ViVaDiag^TM^, RT-PCR and CLIA related to health workers with positive ViVaDiagTM results at the first round of monitoring

## RESULTS

### First Round

94% of all the health workers of our National Cancer Research Center, IstitutoTumori G. Paolo II, Bari, Italy, entered the study. were enrolled. About 92% of the participants had routine daily contacts with clinical departments.

In 7/606 (1.1%) subjetcs, VivaDiag™ (Table 2) provided results that were not negative (weak or strong staining); in all cases an IgM staining was shown (5 weak, 2 strong reaction), in one of these cases the test described a strong simultaneous IgG/IgM intensity. Three cases, comprised the latter one, were health workerswho reported had had recent contacts with COVID-19 patients. None of them presented clinical symptoms associated with COVID-19 disease. Only one was younger than 55 years of age. The day after their VivaDiag^TM^ test, oropharyngeal swabs were collected from all 7 subjects for SARS-CoV-2 RT-PCR testing. None resulted positive for the virus presence (Table 2).

In order to gain further insights into theIgG/IgM analytical sensitivity provided by the colorimetric VivaDiag^TM^kit, a blood plasma aliquot of all 7 subjects was utilized for CLIA analysis of IgG/IgM (Table 2). CLIA showed IgM positivity in three cases (2 nurses and a member of the hospital’s cleaning staff) and the confirmation of strong positivity for IgG/IgM in the third one.

### Second Round

7/393 (1.8%) subjects (Table 2) provided results that were not negative (weak or strong staining): 6 for IgM and 4 for IgG color reaction. In one case a strong reaction was present for IgM and IgG contemporally. The day after the VivaDiag^TM^ test, oropharyngeal swabs was reepeated in all subjects for SARS-CoV-2 RT-PCR testing. One asymptomatic not coming from quarantine, with weak colrimetric reaction for IgM and IgG showed the repesnce of SARS-CoV infection at RT-PCR (Table 2). The same operator resulted positive for IgG expression at CLIA also.

### Monitoring Immunoglobulins

The availability of bllod samples at 2 weeks distance permitted to put in evidence immunoglobulins variations along the time. In summary, after 2 weeks it is observed a shift towards IgG positivity. Becoming not-negative in 4/9 subjects in second sampling versus 1/9 in the first one. Moreover, two cases originally negative showed a weak positivity for both Immunoglobulins at 14 days.

### Interassay comparison

All subjetcs with non-negative Viva-Diag test underwent RT-PCR assay on swab for SARS-COV2 individualization. One 31 yrs old asymptomatic cleaning men with weak positivity for IgM/IgG at Viva-Diag showed the presence of the virus. When Viva-Diag and CLIA assays were compared, CLIA resulted IgM or IgM positive in 4/13 and 4/5 not-negative at Viva-Diag, respectively.

## DISCUSSION

Our study was based on the evidence that nosocomial transmission is a well-known amplifier factor in epidemics diseases and, as a consequence, to try to respond to the need to take care of health of our workers; to permit the rapid identification and isolation of infected HCW, eventually; to protect patients from a possible cross-infection HCW-mediated.

Based on these considerations, we verified the possibility to screen asymptomatic health-care workers with a serological test analyzing IgM/IgG for SARS-CoV2. The study was designed to monitor the kinetics of immunoglobulins response to eventual SARS-COV2 contact and to confirm performance of our colorimetric serological test with respect to standard RT-PCR on swab and CLIA-serologic test.

The first comment concerns the prevalence of serological test-positivity among our asymptomatic HCWs. SARS-COV2 status of HCWs in COVID19 hospitals have been already analyzed whose results recently motivated HCV screening programs by NHS (11). We found 1.3% of our HCW with serological test not negative but all resulted negative for SARS-COV2 test. As expected, this prevalence value is significantly lower than that reported in symptomatic HCWs from Wisconsin (7) and in operating in COVID19 structures experience (12). As far as we know, this is the only study considering asymptomatic HCWs from a non-Covid19 cancer Institute in which, however, a recent activated screening program for all hospitalized patients reported 1% of patients positive for SARS-COV2 (promptly transferred to specific COVID19 hospitals). The prevalence of Ig positive subjects increases to 1.8% in the second round of serological tests (after 14 days) but, interestingly, one of them, (completely asymptomatic) had a successive RT-PCR test positive for SARS-COV2 infection.

The second point of our study concerns the kinetics of immunoglobulins verified through serological test in 393 subjetcs who underwent two blood sampling at 2 wks interval. In general, we observed an increase in IgG positivity in second samples (from 1 to 4 positive cases) and this is in agreement with the common view describing IgM as responsabile of early and IgG late anticorpal reaction (10). In our previous reports (13–14), analyzing serological test with respect to RT-PCR certified virus-infection, we confirmed the early appearance of IgM. Recent data from Guo (10) stressed the variability of Ig kinetics in sARS-COV2 patients showing that IgG specific can be found since early beginning of symptoms and, conversely, that IgM can persist at high level even up to 25 days after symptoms appearance. Our study based on the utilization of a colorimetric qualitative serological test has not been designed to acquire quantitative information on Ig kinetics but it has demonstrated that this assay can be of help in individualization of asymptomatic subjects with supposed occurred contact with SARS-COV virus.

The last point is referred to comparison of Viva-Diag^TM^ with results of molecular RT-PCR assay and CLIA serological test. Noteworthy, Viva-Diag was able to find an asymptomatic HCWs who successively resulted positive at molecular study. This result is important but we have to comment that we screened about a thousand of HCWs (first and second round) to find the positive one, and, even more interesting, that the endpoint of serological tests is not to find the SARS-COV2 positive subjetcs but rather to select those with supposed immunization due to previous contact with the virus (15). For what concerns the comparison of Viva-Diag with respect to qualitative-semiquantitative CLIA analysis of IgM/IgG, our comments concern 41 double performed tests: 23 paired comparisons were performed in quarantine cases HCW coming back from quarantine all resulting negative for IgM/IgG at both tests; the remaining ones were performed in blood samples not-negative at Viva-Diag. CLIA confirmed a positive level of IgM and/or IgG in only 7 out of 18 positive Viva-Diag cases. Conversely, IgG was confirmed positive at CLIA in 4/5 cases positive at Viva-Diag. These results pose the question of different performances of the two tests. It seems CLIA could be less sensitive test in analyzing IgM/IgG presence. However, all people tested not-negative to Viva-Diag were asymptomatic before and after the test, thus leading to the hypothesis of a false positive result of Viva-diag. This impression is confirmed in a further specific analysis we conducted on a larger number of samples paired tested (16). That false positives tests could undermine utility of SARS-COV2 serology testing has been already stressed and has been related with low prevalence of COVID19 in asymptomatic population (17). In fact, prevalence is the key issue conditioning the positive predictive value of any diagnostic test (18).

## Conclusions

Our study suggest that Viva-Diag assay can be of help in individualizing SARS-CoV2 infected people first of all in cohorts of subjetcs with high prevalence of infection. Different performances of serological colorimetric and CLIA tools remain to be further analyzed in larger and specifically designed studies.

## Data Availability

N/A

